# Genetic analyses on the health impacts of testosterone highlight effects on female-specific diseases and sex differences

**DOI:** 10.1101/2021.04.23.21255981

**Authors:** Jaakko T. Leinonen, Nina Mars, Leevi E. Lehtonen, Ari Ahola-Olli, Sanni Ruotsalainen, Terho Lehtimäki, Mika Kähönen, Olli Raitakari, FinnGen, Mark Daly, Tiinamaija Tuomi, Samuli Ripatti, Matti Pirinen, Taru Tukiainen

## Abstract

Testosterone (T) is linked with diverse characteristics of human health, yet, whether these associations reflect correlation or causation remains debated. Here, we provide a broad perspective on the role of T on complex diseases in both sexes leveraging genetic and health registry data from the UK Biobank and FinnGen (total N=625,650).

We find genetically predicted T affects sex-biased and sex-specific traits, with a particularly pronounced impact on female reproductive health. We show T levels are intricately involved in metabolism, sharing many associations with sex hormone binding globulin (SHBG), but report lack of direct causality behind most of these associations. Across other disease domains, including behavior, we find little evidence for a significant contribution from normal variation in T levels. Highlighting T’s unique biology, we show T associates with antagonistic effects on stroke risk and reproduction in males and females.

Overall, we underscore the involvement of T in both male and female health, and the complex mechanisms linking T levels to disease risk and sex differences.

## Introduction

Testosterone (T) is the male sex hormone responsible for regulation of development of primary and secondary male sexual characteristics. Individual variation in T levels has been suggested to shape human physiology broadly, including effects on disease risk in both males and females (1-3). Epidemiological studies and randomized clinical trials for T replacement therapy have observed associations between serum T levels and various traits ranging from type 2 diabetes (T2D) and cardiovascular disease to body composition and behavior (1-11). Yet, these studies have yielded partly mixed results, and, in many instances, the proposed relationships between T, complex traits and disease remain elusive (2, 7-11).

Besides the disease links, T is a known driver for sex differences. After puberty, males and females differ extensively with respect to their average T levels, with males showing roughly 7-15-fold higher serum total T concentrations (1, 12). This difference largely results from the testicular T production in males that far exceeds the amount of T produced in the ovaries and the adrenal gland in females, and is known to directly contribute to variation in, for instance, body composition between the sexes (1, 12).

In the human body, the majority of T is bound to a carrier molecule, whereas only a small fraction (1-3%) of this total T exists as free T, considered to represent the most potent form of T in terms of biological activity (13-15). Most of the remaining T in circulation is tightly bound by sex hormone-binding globulin (SHBG), and the bulk of the rest remains attached to carrier proteins like serum albumin (13, 14). Non-SHBG bound T is often approximated with free androgen index (FAI) (Figure 1A) (4, 13, 14).

**Figure 1.**
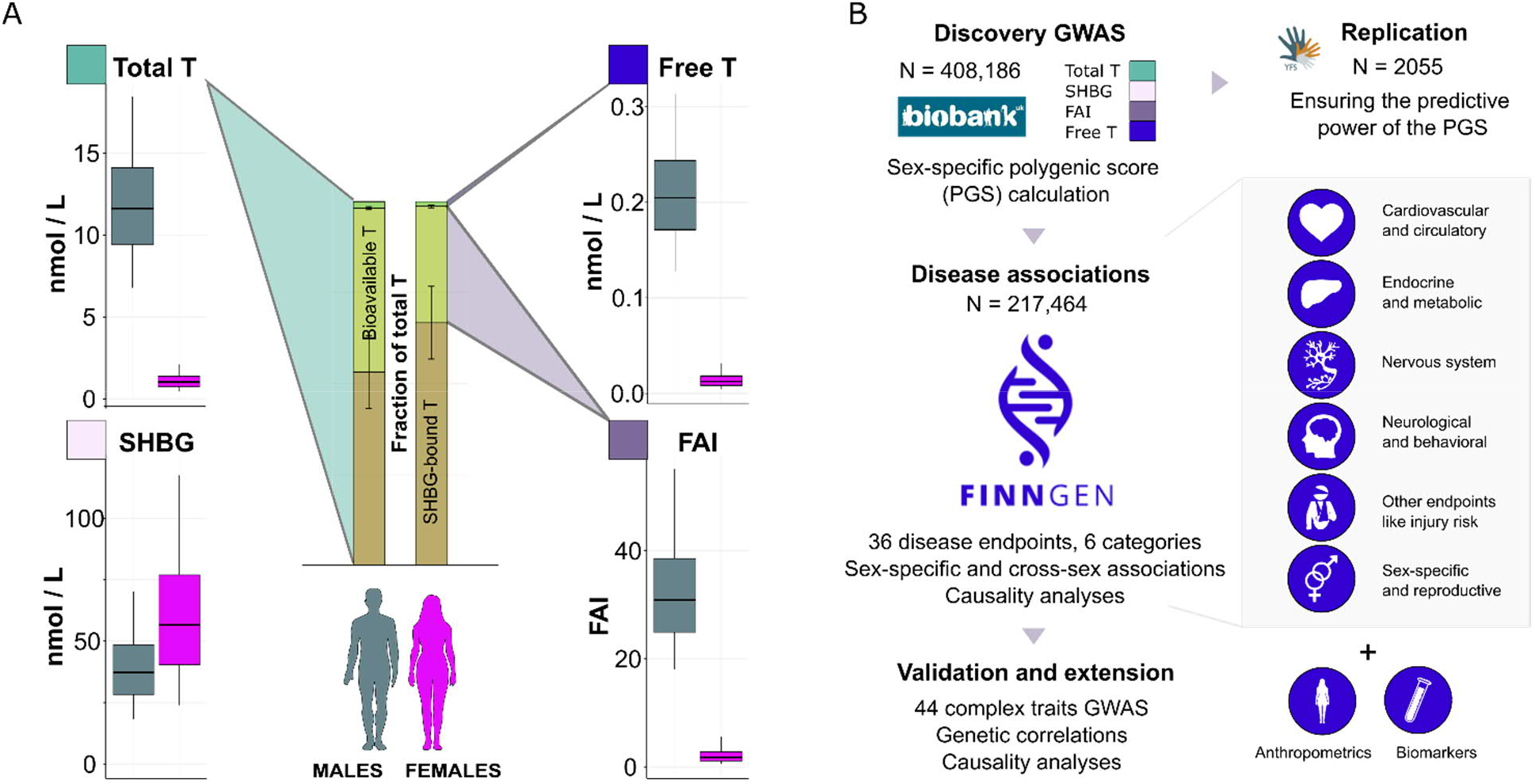
Illustration of the studied traits and study overview. A) Sex-specific distributions for serum total T and SHBG and calculated FAI and free T levels for the UK Biobank participants included in the genome-wide association study (GWAS). The box plots show median (black line), lower and upper quartiles (colored area of the box) and the error bars indicate 5% and 95% quantiles. B) Overview of the study design to assess the contribution of T to health and disease using genetic approaches and biobank data. We conducted the discovery GWAS in the UK Biobank, built sex-specific PGSs for the four T-related traits, validated the PGSs in the Young Finns Study (YFS), and performed complex disease and trait associations in FinnGen (release 5) and using publicly available GWAS data.

Complex physiological processes regulate circulating T levels, allowing T levels to fluctuate based on internal and external stimuli on a daily basis (16, 17). However, twin studies indicate that heritability of serum T is relatively high in both sexes, up to 65% in males (18, 19). Until recently, few genetic variants were shown to associate with circulating T levels and related traits in the general population (4, 18, 20, 21). Yet, through the emergence of genetic and biomarker data from large biobanks, currently more than a hundred loci for serum total T, SHBG and bioavailable T (i.e. free T and FAI) have been identified, with evidence for sex-specific genetic effects (22-24).

Recent efforts have used genetics to address the potential causal contribution of adult T to selected complex traits in both sexes, including T2D, body composition and hormonal cancers, or have examined the effects of free T more broadly in males based on the UK Biobank data (22, 23, 25-27). Here, we extend these investigations combining data from the UK Biobank (N=408,186) and FinnGen (N=217,464). Leveraging the extensive healthcare registry data of FinnGen, sex-specific polygenic scores (PGS), and public data from genome-wide association studies (GWAS), we studied the role of T in both sexes across traits ranging from metabolic conditions and sex-specific reproductive disorders to neurological and behavioral endpoints.

Combining multiple analysis strategies, we illustrate causal links between T and sex-specific diseases and sex-biased traits, including many reproductive conditions in females, while finding that the association between T and many metabolic phenotypes may instead largely stem from shared etiology. Overall, our data provides cues into the biology of T and its contribution to disease risk and complex traits, stressing the unique genetic architecture of serum T levels, and emphasizing the crucial role of the hormone also for women’s health.

## Results

We utilized the rich biochemical and health information available in two population-scale genetic datasets and analysis methods building on GWAS discovery (Figure 1). In brief, we conducted sex-stratified GWAS for T, SHBG, FAI and free T, using data available in the UK Biobank (Figure 1A), from which we built sex-specific PGS for these four traits. The PGSs capture the combined genetic effects on T and SHBG levels, and therefore serve as a proxy for cumulative post-pubertal T exposure. Using an external dataset (Young Finns Study; YFS), we validated the performance of the PGSs. We then investigated the effects of the PGSs on a wide range of diseases across diverse clinical entities using the FinnGen study (Figure 1B). Lastly, we evaluated causal relationships and genetic correlations between the studied T traits and complex traits, leveraging publicly available GWAS summary statistics.

### GWAS and polygenic scores for testosterone traits

We identified more than a hundred genome-wide significant (*p*<5e-08) loci for all testosterone traits (up to 263 loci for SHBG in males) in the UK Biobank GWAS (Methods; Supplementary Tables 1-9). In both sexes, common variants (allele frequency >1%) covered a large proportion of the trait variability, with the SNP heritability (h^2^) estimates ranging from 10% for total T in females to 28% for SHBG in males (Supplementary Table 9). The associated loci were enriched for genes affecting steroid hormone biosynthesis, metabolism and excretion, with preferential expression in the liver for all the studied traits, in line with recent findings (Supplementary Figures 1&2, Supplementary Table 10) (22-24).

The loci affecting SHBG were largely shared between the sexes (genetic correlation (r_g_) =0.88, *p*=9.7e-197), for FAI sharing was intermediate (r_g_=0.54, *p*=5.8e-26), but we observed a near-zero genetic correlation estimates for both serum T and free T between males and females (r_g_=0.08 and 0.05, respectively, *p*>0.05), indicating sex-specific genetic determinants, as previously reported (22, 23). Co-localization analyses between the male and female GWAS further confirmed the widespread sex-specificity of the genetic loci (Methods, Supplementary Figure 3 and Supplementary Tables 11-19) (23).

Reflecting the sex-specific genetic architecture, we observed strong genetic correlation between total T and SHBG only in males (r_g_=0.78 in males vs. 0.05 in females, Supplementary Figure 3 and Supplementary Table 9). In females, instead, the genetic determinants for FAI and free T were shared with SHBG (e.g. r_g=_-0.80 with FAI). Highlighting these connections, SHBG was found causal for total T levels in males (genetic causality proportion (GCP)=0.80, *p=*5.8e-05, Methods), whereas in females SHBG appeared to control especially FAI and free T fractions (GCP=0.83, *p=*3.8e-07 for free T, Supplementary Table 9).

To study the impacts of T in datasets where T measurements are not directly available, we next constructed sex-specific genetic predictors for T levels, PGS, for each trait applying the LDpred algorithm (28) to the sex-specific GWASs (Methods). We tested the predictive ability of the PGS in the YFS where the phenotypic variance explained by the PGS (R^2^) ranged between 1.1% (male free T) to 9.2% (male SHBG), indicating the PGS predict T and SHBG levels in an independent cohort (Supplementary Table 20). Notably, the sex-specific PGSs for T and free T had no predictive value in the opposite sex (Supplementary Table 20 and Supplementary Figure 4).

### Studying the links between cumulative T exposure and disease

We continued by using the PGS to study how post-pubertal T exposure associates with disease risk, the associations potentially implying causal relationships (28). To this end we used the FinnGen data, consisting of 217,464 (94,478 males, 122,986 females) Finnish participants, representing roughly 5% of the Finnish adult population, with genotypes linked to up to 46 years of follow-up within nationwide healthcare registries (29). We studied 36 diseases with potential links to hormones from the following categories: 1) endocrine and metabolic problems, 2) sex-specific endpoints (many specific to females, e.g. postmenopausal bleeding, (PMB)), 3) cardiovascular and circulatory system, 4) nervous system disease, 5) behavioral and neurological diagnoses and 6) other endpoints like injury risk (Supplementary Table 21). The number of cases ranged from 229 individuals diagnosed with hirsutism to 68,774 statin users.

We first tested whether the PGS are associated with disease risk in a sex-specific manner, observing 32 associations (*p<*0.0014, after Bonferroni correction for 36 independent tests; Supplementary Table 21 and Supplementary Figure 5). The associations involved mainly endocrine, metabolic and sex-specific disorders, highlighting in particular female-specific endpoints (Figure 2A&B, and Supplementary Table 21). In males, both total T and SHBG PGSs often associated with reduced disease risk, whereas we saw few associations to free T. In females, higher total T, FAI and free T PGSs generally increased risk for multiple diseases, often showing inverse associations to SHBG. Given the shared association profile with SHBG, we additionally included the SHBG PGS as a covariate in the analyses to distinguish true T-driven effects.

**Figure 2.**
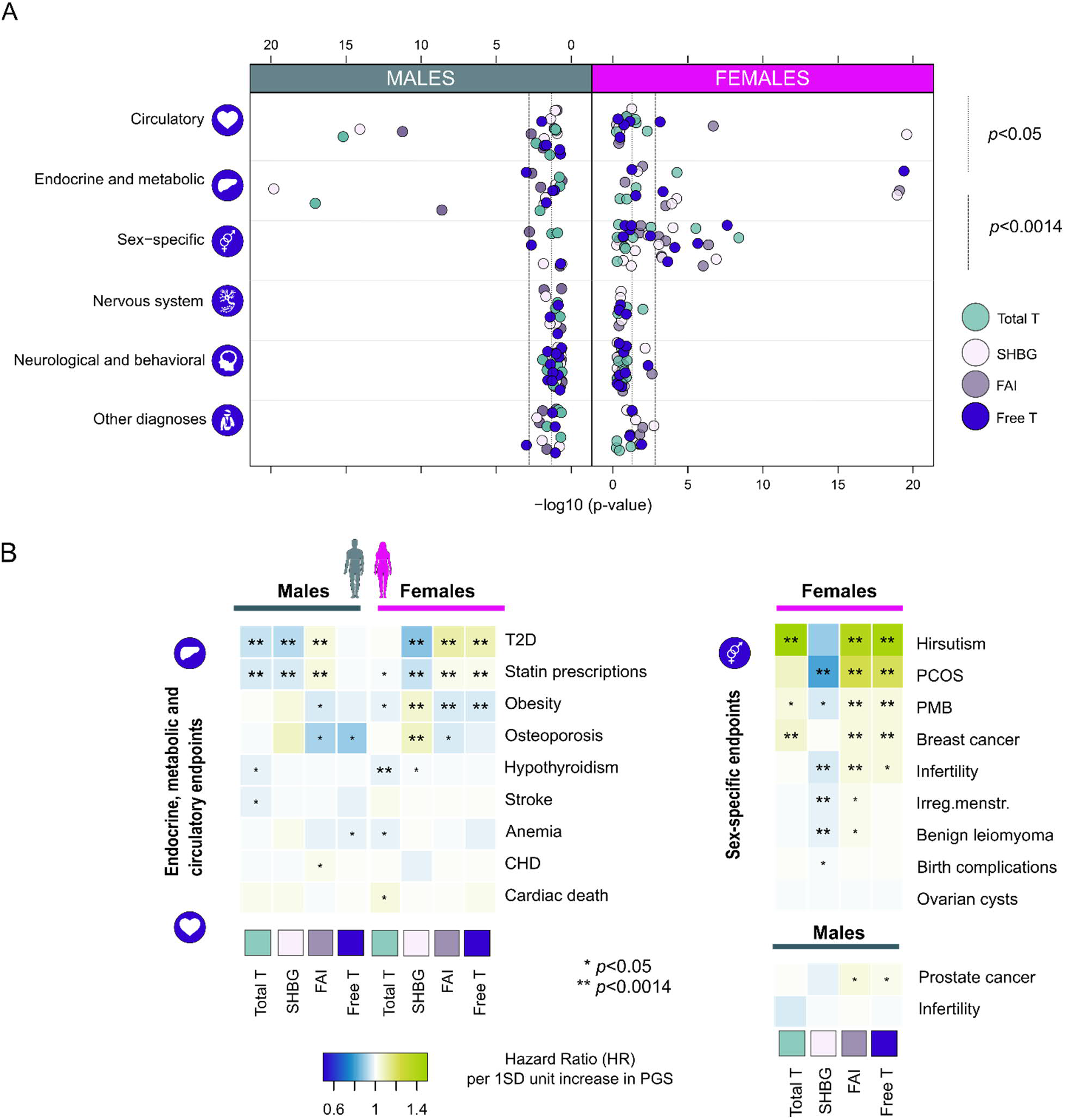
Results from the PGS associations with disease endpoints in the FinnGen. Panel A illustrates the distribution of association *p*-values by disease category for each biomarker PGS separately for both sexes. For clarity *p*-values capped at 1e-20. Panel B shows hazard ratios per one SD increase in PGS for 20 traits from endocrine, metabolic, circulatory and sex-specific categories. \**p*<0.05, \*\**p*<0.0014, corresponding to Bonferroni correction for 36 independent traits. PCOS = polycystic ovary syndrome, PMB = post-menopausal bleeding, CHD = coronary heart disease.

Underscoring T’s and SHBG’s involvement in metabolism, larger PGS values for total T and SHBG were associated with reduced T2D risk and statin use in males (for total T in males, HR=0.94, *p=*1.4e-17 and 0.96, *p=*1.1e-15, respectively). SHBG appeared protective of these endpoints also in females (HR=0.87, *p=*4.8e-58 and 0.95, *p=*1.1e-21), whereas higher female-specific free T PGS increased risk for both (HR=1.09, *p*=1.9e-22 and HR=1.02, *p*=0.0011). The SHBG adjusted analyses nevertheless suggested the T and free T associations to these metabolic traits in either of the sexes were not primarily attributable to androgen action (Supplementary Table 21 and Supplementary Figure 6). In females, higher T PGS was additionally associated with lower hypothyroidism risk (HR=0.97, *p*=6.5e-05), persisting SHBG adjustment (HR=0.97, p=8.6e-05). We also detected suggestive associations to bone strength and injury risk in both sexes, but with the exception of SHBG and osteoporosis in females (HR=1.08, *p=*0.00015), none of these findings survived correction for multiple testing.

In the sex-specific category, we replicated the known associations of T to PCOS and breast cancer risk in females (22) (HR=1.02, *p*=2.8e-06 and HR=1.04, *p*=0.0001 for free T PGS) (Figure 2B and Supplementary Table 20). Of the novel endpoints, we robustly linked T with hirsutism and post-menopausal bleeding (HR=1.45, *p*=2.7e-08 and HR=1.05, *p*=0.00032 for free T PGS). The association to hirsutism (excessive hair growth in a male-type fashion) appeared particularly pronounced, with the risk of the condition almost doubling with a 2SD change in free T. With the exception of PCOS, total T and free T displayed similar associations to these traits. The SHBG adjusted analyses suggested these associations were androgen dependent (Supplementary Table 21 and Supplementary Figure 6). Whereas SHBG adjustment generally showed negligible effects on total T associations, for free T and FAI, linked also with infertility risk in females (HR=1.04, *p=*0.00076 for FAI), some confounding by SHBG (showing generally positive effects on reproductive traits, e.g., HR=0.98, *p=*0.00116 for irregular menstruation) was evident.

We observed no statistically significant associations to other diseases (Figure 2A). We for example detected no associations to any of the 13 neurological/behavioral endpoints studied, including Alzheimer’s disease, alcohol use, and anxiety disorders (all *p*>0.0014) in both sexes. However, the PGSs did show some nominal clinically-relevant associations, e.g., higher PGSs for male free T was linked with prostate cancer and lower osteoporosis incidence (HR=1.03, *p*=0.0083 and HR=0.89, *p*=0.0023, respectively). Male total T associated with stroke (HR=0.97, *p=*0.017), and female total T with cardiac death (HR=1.04, *p=*0.049) and Parkinson’s (HR=1.08, *p*=0.017). Both male free T and female total T showed associations to lower anemia risk (HR=0.97, *p*=0.042 HR=0.98, *p*=0.041, respectively, Figure 2, and Supplementary Table 21).

### Evaluation of causal relationships between T and disease

Although the PGS associations may imply direct causality of the first trait to another, these are also prone for confounders including genetic pleiotropy (30). We estimated causal relationships between T and the studied endpoints using two complementary MR methods that correct for potential pleiotropy: LCV (31), representing a genome-wide approach, and MR-Egger (32) that uses significantly associated SNPs. For the sex-specific PGS data, in 23/252 instances one or both methods suggested evidence for a causal relationship (*p*<0.0014) between a PGS and a disease (Figure 3, Supplementary Figure 7, and Supplementary Table 22). Finally, we distinguished between the effects of T and SHBG by including the latter as a covariate in multivariable MR Egger analyses (33).

**Figure 3.**
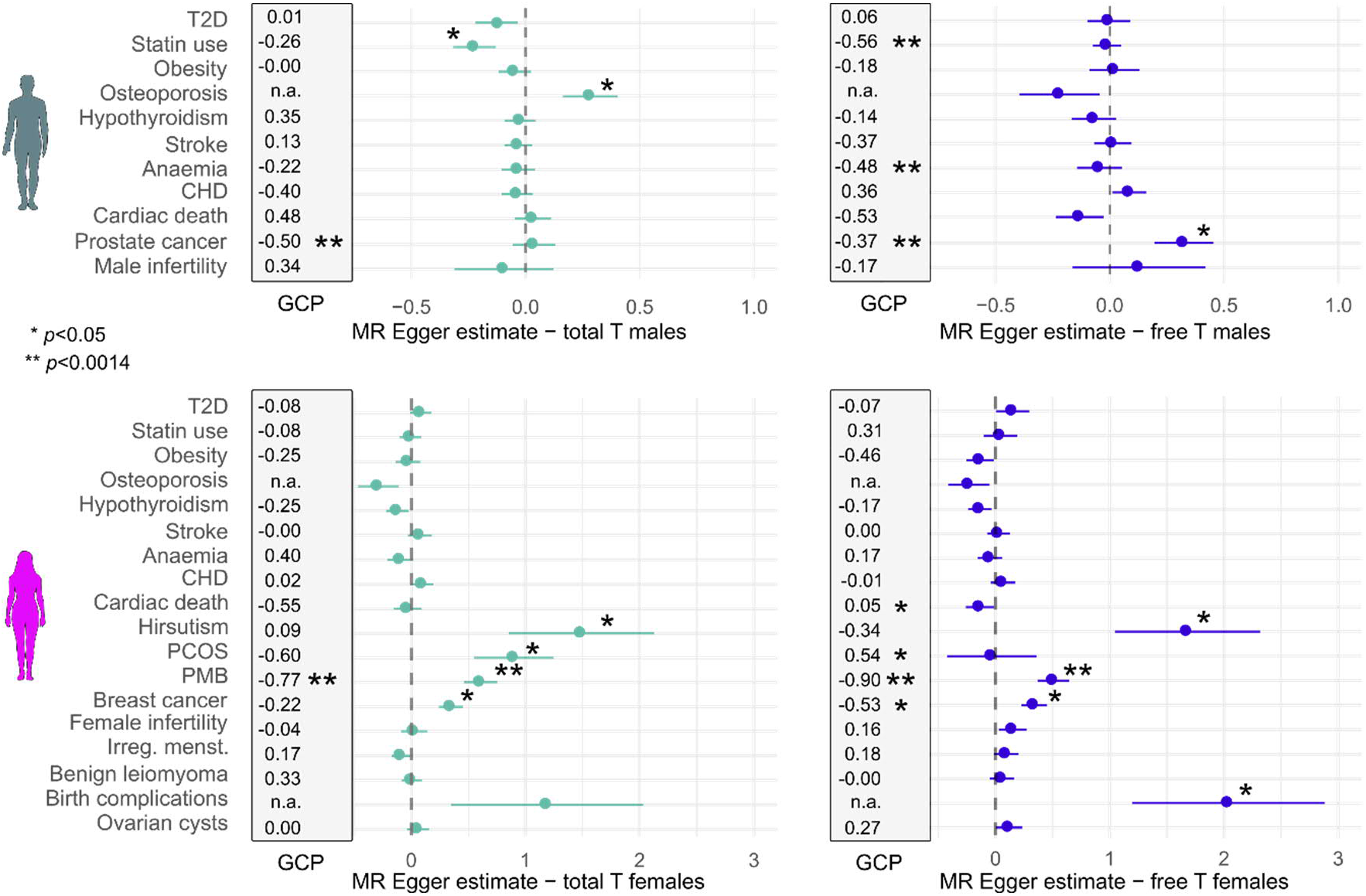
Results from the causality analyses in FinnGen, showing MR estimates for total and free T in males and females. The figure includes traits from the endocrine, metabolic, circulatory and sex-specific categories shown in Figure 2. Shown are both genetic causality proportion (GCP) estimates from LCV (within the grey boxes, 0=no causality, 1= fully causal, minus sign indicates suggested reverse causality i.e. the endpoint affecting T), and MR Egger betas and standard error for each trait (horizontal lines). The MR Egger value in the x-axis corresponds to SD increase in risk/per SD increase in T/free T. \**p*<0.05, \*\**p*<0.0014, n.a.=not applicable due to low heritability estimate in FinnGen under the LCV model.

Despite the many associations (Figure 2), we observed no clear evidence for T being directly causal for metabolic traits. For example, the link between total T, osteoporosis and statins in males appeared confounded by SHBG (the effect being close to zero in the adjusted model, (Figure 3 and Supplementary Table 22)). We also consistently saw no significant causality between T and T2D, including both sex-combined and sex-specific T2D GWASs from FinnGen (Figure 3, Supplementary Figure 7 and Supplementary Table 22).

In contrast, the causality analyses supported the role of T in the regulation of female reproductive health. Adjusting for the effects of SHBG in the multivariable MR Egger analyses seemed to strengthen these causality estimates (Supplementary Table 22). Under both MR Egger models, we observed causality between total and free T and postmenopausal bleeding and hirsutism (β=0.61, *p*=4.5e-05 and β=2.11, *p*=0.002, respectively, for free T in multivariable MR Egger). For PCOS MR Egger supported causality for total T (β=0.90, *p*=0.012) and LCV for free T (GCP=0.54, *p*=0.0017) (Figure 3 and Supplementary Table 22). Additionally, both approaches indicated causality between T and hormonal cancers (e.g. GCP=-0.55, *p*=0.041 and β=0.34, *p=*0.0031 for female free T and breast cancer; GCP=-0.37, *p*=2.8e-16; β=0.32, *p*=0.014 for male free T and prostate cancer). Notably, genetically predicted free T was linked with increased cancer risk also in the opposite sex (GCP=-0.37, *p*=6.6e-19 for male free T and breast cancer, and GCP=-0.74, *p*=0.0041 for female free T and prostate cancer)..

Lastly, some causal relationships were detected despite no robust associations in the original PGS analyses. For example, T seemed to protect females from seropositive rheumatoid arthritis (RA) (Multivariable MR Egger β=-0.48, *p*=0.0025 for free T), and T and SHBG were linked with injury risk in both sexes (e.g. GCP=0.53, *p*=0.0018 and β=0.17, *p*=1.3e-05 for SHBG increasing forearm/elbow injuries in males). Finally, LCV suggested a causal relationship between higher SHBG and ADHD in both sexes, and higher free T and increased risk for conduct disorder but decreased risk for emotionally unstable personality in males (*p*<0.0014, Supplementary Table 22), pointing to potential hormonal involvement in the regulation of neuronal processes.

### Results from the cross-sex PGS associations in FinnGen

Owing to the unique genetic architecture, the sex-specific PGS for T and free T do not predict the corresponding hormone levels in the opposite sex (Supplementary Table 20). Given this, we reasoned that cross-sex analyses, i.e., analysis of the effect of a sex-specific PGS in the opposite sex, would provide us with an additional means to assess if the original associations stem from T action, and to detect potential antagonistic effects for the PGSs between the sexes.

Aligning with the results from the MR analyses, 15/20 of nominally significant (*p*<0.05) sex-specific associations remained similar (Z-test *p*>0.05 for difference in PGS effects) also in the cross-sex analyses, pointing to shared genetic etiology rather than T action as the basis for many metabolic associations. For example, both female and male total T PGSs, with and without SHBG adjustment, associated with hypothyroidism risk with a similar effect size in the opposite sex, and all PGS associations to T2D were replicated in the other sex (Figure 4A, and Supplementary Tables 23&24).

**Figure 4.**
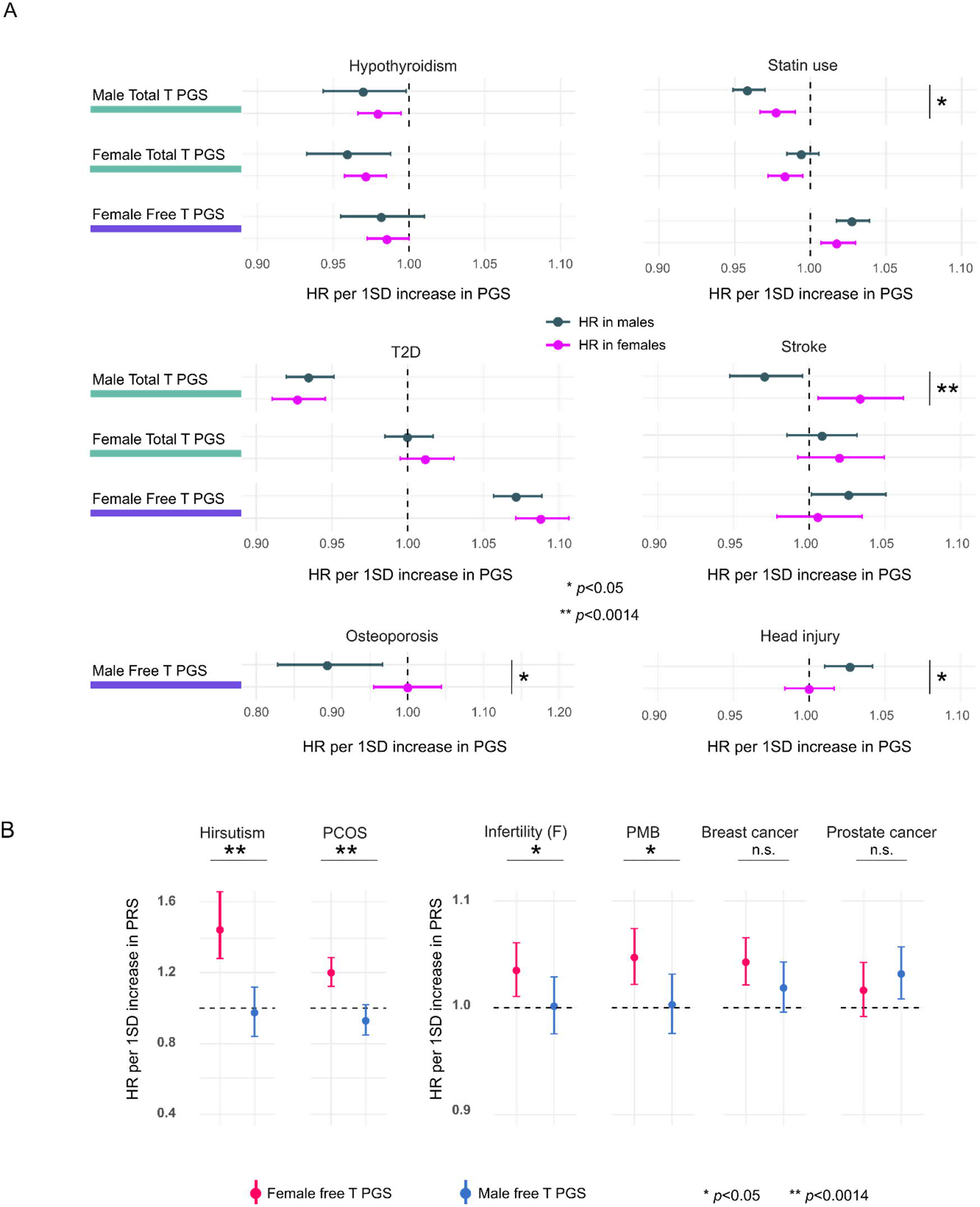
Results from the cross-sex analyses. **A)** Cross-sex PGS associations with hypothyroidism, statin use, T2D and stroke. *p*<0.05 suggests that the effects of a given PGS vary depending on sex **B)** Illustration of cases with statistical evidence for male and female PGSs having different effects (hirsutism, polycystic ovary syndrome (PCOS), infertility and post-menopausal bleeding (PMB). No evidence for such difference for breast and prostate cancers.*Chi-Squared *p*<0.05, ** = Chi-Squared *p*<0.0014, n.s. = not significant. Panels A and B show HR point estimates with 95% confidence intervals.

The associations differed (Z-test p<0.05) in the opposite sex for male total T PGS and stroke, female total T PGS and anemia, and male free T PGS, head injury risk and osteoporosis. (Figure 4A, and Supplementary Tables 23&24). In addition, although an effect of male total T PGS on statin use was observed also for females, it was attenuated. Intriguingly, while higher male total T PGSs reduced stroke risk in males (HR=0.97, *p*=0.017), this associated with increased risk for stroke in females (HR=1.03, *p*=0.017). This was the only endpoint for which we detected evidence for sexual antagonism. The antagonistic effect enhanced with SHBG adjustment (HR=0.96, *p*=0.011 in males and HR=1.07, *p*=0.0010 in females). The results indicate the genetic effects on stroke risk may be partly sex-specific (34), with potential interplay from sex hormones.

Finally, the cross-sex analyses implied increased androgen load as a direct contributor to poorer reproductive health in females, agreeing with the MR-based causality assessments. For the reproductive endpoints that were associated with female free T PGS, i.e., infertility, PMB, PCOS and hirsutism, the male free T PGS had no predictive power (all *p*<0.05, Figure 4B, Supplementary Tables 23&24). Nonetheless, the effect sizes were not attenuated in a statistically significant manner for breast and prostate cancers, (*p*>0.05, Figure 4B), echoing the causality analyses and pointing to some degree of shared genetic risk, irrespective of free T levels, between male and female hormonal cancers.

### Extending the FinnGen discoveries

We next sought to validate and refine the FinnGen discoveries in additional datasets, extending our analysis to include quantitative traits not available in large numbers in FinnGen. To this end, we used genetic correlation analysis, allowing for estimation of the extent to which two traits are affected by the same genetic factors (35, 36), followed by causality estimations.

We selected 44 traits with publicly available GWAS summary statistics, identical to (e.g., T2D, breast and prostate cancers) or closely reflecting the studied disease phenotypes (heel bone mineral density (HBMD), mood swings) from FinnGen, adding anthropometric traits to the analyses (Supplementary Table 25). For most female-specific phenotypes studied in FinnGen, including hirsutism and PMB, we had no comparable phenotypes, as there are no published GWAS available.

We found evidence of significant genetic correlation in 72/352 instances (*p<*0.0011, corresponding to Bonferroni correction for 44 independent tests) (Figure 5, Supplementary Table 25) with the results reflecting the FinnGen PGS associations. We observed significant genetic correlations to traits related to metabolism, including many biomarkers and anthropometrics, but detected only few correlations to behavioral traits, and no significant correlations to neurological or temperamental traits (Figure 5, Supplementary Table 25). Notably, in all cases where we observed genetic correlation to behavioral traits, these had clear links to metabolism (smoking, sleep duration and exercise).

**Figure 5.**
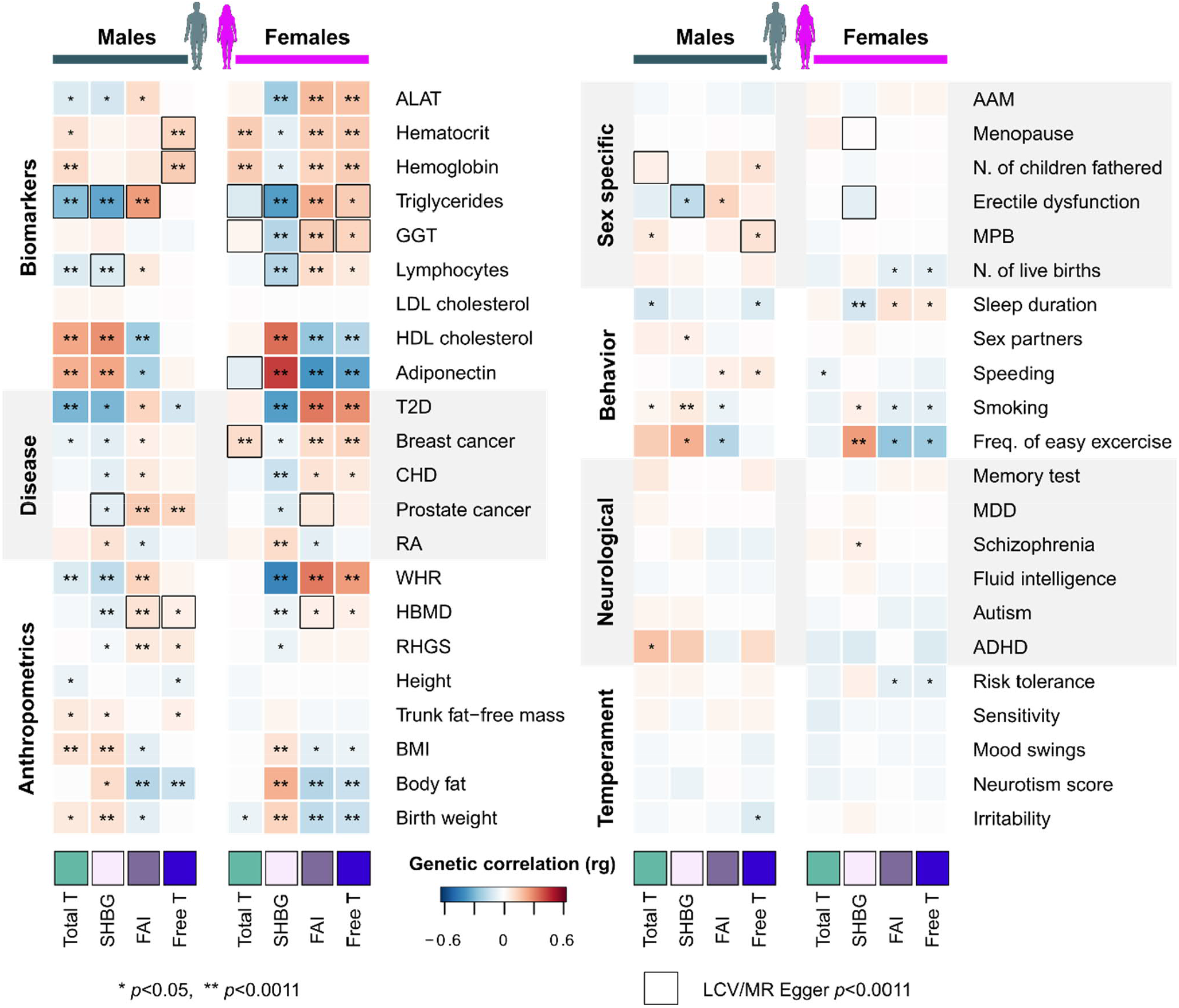
Genetic correlation and causal relationships between the 44 traits and phenotypes from public GWAS data based on LDSC and MR analyses. * *p<*0.05, ** *p<*0.0011 (significant correlation after Bonferroni correction for 44 independent tests). Black boxes = (*p<*0.0011) for a causal relationship between the tested traits based on LCV and MR Egger analyses. AAM = age at menarche, ALAT = alanine transaminase, ADHD = attention deficit and hyperactivity disorder, CHD = Coronary heart disease, GGT = Gamma-glutamyl transferase, HBMD = Heel bone mineral density, MDD = major depressive disorder, MPB = male pattern baldness, RA = rheumatoid arthritis, RHGS = relative handgrip strength, WHR = waist-to-hip ratio.

The genetic factors increasing serum total T and SHBG appeared to promote a favorable metabolic profile in males, supporting the PGS findings. Despite correlating with increased BMI, total T and SHBG were positively correlated to adiponectin, high-density lipoprotein (HDL) and lower waist-to-hip ratio (WHR) (r_g_>0.20, *p<*0.0011) whilst lowering triglycerides and T2D incidence in males (r_g_<-0.25, *p*<0.0011). In contrast, higher FAI and free T fractions in females associated with negative metabolic effects, including higher WHR (r_g_=0.25, *p*=1.6e-22) and lower HDL cholesterol (r_g_=-0.18, *p*=4.2e-05), whilst free T showed no significant correlations to these metabolism-related traits in males. Strong correlations were observed also between SHBG and metabolic traits in females, including negative associations to markers of liver damage (alanine transaminase (ALAT), r_g_=-0.21, *p*=1.7e-08) and gamma-glutamyl transferase (GGT), r_g_=-0.16, *p*=0.0001).

We observed significant genetic correlations to hormonal cancers in both sexes, in line with recent findings (22, 25). In males, the genetic factors increasing FAI and free T promoted prostate cancer (r_g_=0.12, *p*=0.0004), whereas in females these increased especially the risk of estrogen receptor (ER) positive breast cancer (r_g_=0.15, *p*=9.1e-09). Finally, consistent with existing genetic, epidemiological and experimental data (1, 25), the genetic correlation analyses pointed towards shared genetic background for T, hemoglobin levels and body fat in both sexes (e.g. r_g_=0.15, *p*=1.0e-07 and r_g_=-0.14, *p*=0.0002 for hemoglobin and body fat, respectively, with male free T).

In the LCV and MR-Egger analyses we found statistically significant evidence (*p*<0.0011) of a causal relationship in 7% (26/354) of instances across the 44 traits (Figure 5 and Supplementary Table 26). Reflecting the results from FinnGen, despite the genetic correlations to many traits there was no evidence for significant causality, but the few suggested causal relationships involved traits with clear biological links to T function.

As examples of expected causal relationships (1, 25, 37), our analyses supported the contribution of free T levels to male-pattern baldness (MPB, GCP=0.44, *p=*7.7e-19) and hemoglobin levels (GCP=0.64, *p=*0.00085) (Figure 5 and Supplementary Table 26). In addition, despite the lack of significant genetic correlations between these traits, higher T in males was linked to increased number of children fathered (NCF, GCP=0.62, *p=*1.2e-15 for total T, GCP=0.42, *p=*0.011 for free T), raising speculation about the potential evolutionary benefits of males maintaining adequate T levels.

The MR Egger analyses supported the causality of female total T and free T to ER+ breast cancer (β=0.282, *p*=0.00014 and β=0.250, *p*=0.002, respectively). Instead of T, in these analyses prostate cancer risk was linked to SHBG levels in males (GCP=-0.64, *p*=0.00019) (38). Additionally, SHBG increased lymphocyte count in both sexes (GCP=0.39, *p*=1.6e-07 in males, GCP=0.42, *p*=0.00014 in females) (39), and in males, we linked SHBG also with reduced risk of erectile dysfunction (GCP=0.42, *p*=0.00032). In females, we observed causality between SHBG and age at menopause (β=-0.756, *p*=0.0010), suggesting that inherited differences in SHBG levels likely modify reproductive phenotypes across female lifespan.

In most instances, we however found no evidence of a significant causal relationship between T levels and the studied traits (Supplementary Table 26). Yet, emphasizing the intricate relationship between hormone levels and metabolism (40), some connections to metabolism-related biomarkers emerged: for example, it appeared that triglycerides may causally influence T (GCP=-0.48, *p*=1.3e-30) and free T (GCP=-0.80, *p*=1.4e-06) levels in females. In combination with the PGS analyses, these results thus further suggest that whilst some sex-biased phenotypes may be directly related to T levels, in most instances T’s relationship to complex traits and diseases is not straightforward.

## Discussion

Since its discovery in the early 20^th^ century, testosterone (T) has been proposed to modify phenotypes and diseases that differ between the sexes, due to the extensive male-female differences in circulating T levels. Besides the biological impacts of normal variation in T levels, the use of T replacement therapy in medical practice has been globally on rise, and the potential risks and benefits associated with T supplementation remain debated (7). To provide a broad and systematic perspective into the function of T as a regulator of health and disease in males and females we leveraged the UK Biobank resource to construct PGSs - predictors for genetically determined T levels - for both males and females, which we then associated with a uniquely rich collection of disease endpoints from 217,464 participants in FinnGen. This combination allowed us to extend and refine recent efforts that have utilized genetic data to understand the disease impacts of T, concentrating on a limited set of phenotypes or only to males (22, 23, 25-27). In addition, by taking advantage of the sex-specific genetic determinants of T in cross-sex analyses, and through careful Mendelian Randomization strategies, we could pinpoint T’s causal effects on adult health.

Based on our analyses, three major themes emerged regarding T’s contribution to disease. First, we report that the studied PGSs associated with disease risk especially in females. Secondly, we highlight distinct association profiles for total T and free T in males, consistent with proposed divergent biological effects for the bound and unbound T fractions (13-15). Underscoring the potential role of SHBG as a confounder, the former closely correlates with SHBG levels in males, and the latter in females (13-15). Thirdly, taking into account such confounding, we generally observed causal relationships between a genetic predisposition to higher T levels and several sex-specific and sex-biased phenotypes with clear biological links to T, but less contribution to most other phenotypes, echoing experimental data and findings from recent MR studies (22, 25).

Besides the causal links to several female-specific reproductive endpoints, sex-biased traits and hormonal cancers, significant associations involved diseases and traits from the metabolic and endocrine categories. We stress the apparent complexity of these relationships. For many metabolic traits SHBG – either directly or potentially through its action on other hormones - appeared to confound T’s associations and causality estimates. Beyond this T/SHBG relationship, we furthermore estimate that some of the observed associations to metabolic health, including T2D and hypothyroidism, may generally reflect widespread genetic pleiotropy and thus overall complex shared genetic etiology rather than T action.

Based on our data, we further speculate that normal variation in T levels - contrary to popular beliefs - has only modest effects on most phenotypes. Particularly, the grounds to explain some temperamental and neurological phenotypes like anxiety and emotional instability with heritable differences in adult T levels (41, 42) appears unsubstantiated in the light of our study. Although we cannot fully exclude a causal relationship between T and some of these phenotypes, our data suggests that especially without larger sample sizes or refined phenotyping, efforts to link T levels to behavior will likely be unproductive. Taken together, supporting recent recommendations, our data thus suggests that the risks and benefits of using T as a medical treatment should be carefully weighted, given T’s complex and indirect relationship to most phenotypes and potential adverse and beneficial outcomes in both sexes (7, 25).

Having comprehensively mapped the impacts of T across diverse complex disease and traits, we can start drawing inferences on the role of T as a contributor to the male-female differences. Indeed, in such instances where causality of T was implicated for a sex-shared trait, the effect estimates often aligned with the direction of the phenotypic sex bias in the given trait, i.e., higher T levels associated with typical male characteristics. For instance, our work highlighted causal connections between increasing free T, higher hemoglobin and higher bone strength, backed up by previous experimental observations (1, 43, 44). Extrapolating from the MR results, we estimate that ∼10-20% of the mean difference in hemoglobin levels between males and females may result from average differences in free T levels, consistent with the notion that T directly affects male-female differences in, e.g., athletic capacity (1). Moreover, higher free T levels were found causal to masculine external features like hirsutism and baldness, and in females higher bioavailable T levels correlated genetically with a shift of metabolism into a male-like direction (e.g., increased BMI, WHR, and poorer blood lipids, yet reduced risk for obesity and reduced body fat levels).

Overall, the limited evidence for the causal involvement of T for most of the studied traits however suggests that most phenotypic sex differences are not attributable to a linear relationship between T levels and a phenotype. Instead the impact of T can be mediated through a threshold effect, potentially at a given developmental time window, acting as a switch that results in more global rewiring of biological processes and thereby in systematic male-female differences. Under this model, the within-sex variability in T may have non-existent or very subtle effects on many phenotypes, complicating the detection of potential causality of T.

Finally, our study provides some unique insight into the potential causes behind the sex differences in T levels. In the cross-sex PGS analyses, we found only one case where there was clear evidence of antagonistic effects for the T PGS between the sexes. The male-specific total T PGS protected from stroke in males, whereas the same PGS had opposite effects on stroke risk in females. Although we cannot conclude that the action of T truly drives these associations, the result agrees with a degree of sex-specificity in the genetic disease mechanisms for stroke (34).

The observation that the genetic variants responsible for regulating T levels are largely distinct between males and females moreover raises speculation about the evolutionary forces maintaining sex differences and shaping the biology of T. The cross-sex genetic correlations for traits related to fitness (e.g., reproductive success) are generally expected to be low, due to potentially conflicting evolutionary pressures (45). We indeed associated T positively with reproductive success in males (T increasing the number of children fathered), but negatively with both pre- and postmenopausal reproductive health in females with evidence for a causal role of T behind these associations. We may thus speculate that there exists a selective advantage in relation to reproductive success to maintain higher T levels in males, whereas the opposite may be true for females, potentially promoting the widespread sex differences across multiple traits.

Although based on extensive data sets, our study still has some limitations. We stress that studying T levels differs drastically from studying T action, serum T levels serving only as a proxy for the latter. Furthermore, although we extrapolate that the PGS for total T and SHBG used in the study explain a substantial fraction of the variance in these traits in the studied cohorts, for example the male free T PGS seemed to yield less accurate predictions. Genetic studies are also prone to confounding by pleiotropy, whereby a gene influences multiple traits via independent biological pathways (30). Although we opted for adjusting for the effect of both SHBG and BMI in our analyses – both known confounders for testosterone levels (Supplementary Figure 8) (46, 47) - it remains possible that these connections still affect our findings. Genetic pleiotropy may confound also MR analyses, and despite the vast potential of MR in establishing causal relationships (48), we generally propose caution in interpreting these findings. In our case the MR models did not always agree on causality (Supplementary Tables 22 and 27), common to recent MR studies assessing the function of T (22, 23, 25). When based on large number of variants, the MR analyses often include variants that do not fulfill the strict definition of an instrumental variable. Here we show via MR and cross-sex PGS analyses that such pleiotropic variants may underlie for example T’s associations with T2D and hypothyroidism.

Importantly, our setting does not allow for assessing the effects of fetal T exposure, which may be critical, e.g., for neurological traits (49). We also emphasize that our results are based on normal variation in T levels, not on supraphysiological T injections. Additionally, many of T’s effects depend on its conversion to estradiol also in males, and we cannot rule this out as a potential confounder in our study. Finally, the data used in our study does not allow for assessing the effects of acute changes in hormone secretion, and personal differences in the response to such fluctuations may be crucial for some phenotypes.

Despite these challenges, we were able to highlight several novel albeit often expected relationships with genetically determined T levels, human health, and sex differences. Besides the gained medical insight, underscoring some critical factors that should be considered when assessing these relationships, with this study we thus provide a reference point for future genetic and epidemiological studies studying the action of T.

## Methods

### Genotype and phenotype data from the UK Biobank

The genetic association analysis was based on data from the UK Biobank, a population-based biobank consisting of 502 637 subjects (aged 37-73 years)(50). At recruitment, participants provided electronic signed consent. Ethics approval for the UK Biobank study was obtained from the North West Centre for Research Ethics Committee (11/NW/0382). All experiments were performed in accordance to relevant guidelines and regulations including the Declaration of Helsinki ethical principles for medical research. This study was run under UK Biobank application number 22627.

Multiple biochemical assays have been performed on the entire UK Biobank cohort, including measurements of serum testosterone, SHBG and serum albumin. These measurements were performed once for each participant for using Beckman Coulter DXI 800 Chemiluminescent Immunoassay, with competitive binding for testosterone and two-step sandwich for SHBG, referenced in https://biobank.ndph.ox.ac.uk/showcase/showcase/docs/serum_biochemistry.pdf. The document contains also details about the extensive QC procedures for all biochemical measurements in the UK Biobank. For example, assay consistency was monitored by using internal QC samples between batches, and external quality assurance schemes against the ISO 17025:2005 standard.

We restricted our study to encompass 408,186 individuals from the white British subset. We removed outliers for genotype heterozygosity and missingness, as well as samples with sex chromosome aneuploidies, mismatches between reported and inferred sex, and samples that UK Biobank did not use in relatedness calculations (50). We did not exclude related samples since our analysis method (BOLT-LMM) allows for their inclusion.

For the GWASs, we used biochemically measured testosterone and SHBG, and calculated FAI and free T. For calculation of FAI, we used the formula *100*Testosterone/SHBG* (nmol/ml). Calculated free T was derived using the Vermeulen equation using directly measured albumin values for each participant in the equation, as described in (51, 52).

All four traits were separated by sex and log transformed. A linear regression model was fit for each trait with BMI and age as covariates, as well as menopause status for females. Subjects with residuals values of +-5 SD from the mean were excluded from the analyses, serving as a further QC step to exclude outliers potentially reflecting medical conditions or drug use affecting androgen levels. Inverse normalized values of the remaining residuals were used as phenotype values for the GWAS analyses. After the QC-steps, our study included altogether 177,499 males and 205,141 females.

### Genetic association analysis and definition of the lead SNPs

The GWAS analyses were performed using BOLT-LMM (v2.3.2)(51, 52). Imputed SNPs were restricted to variants with MAF ≥ 0.1 % and imputation quality ≥ 0.7 (50). 1000 Genomes European data was used as reference LD scores for calibrating the BOLT-LMM statistic. First 10 Principal Components were used as quantitative covariates in the runs. A linear regression model was fit for each trait with BMI and age as covariates, as well as menopause status for females. Genetic correlation analyses, heritability estimates and number of loci found implied the results remained consistent with different covariate configurations (Supplementary Figure 8), but we chose to include body mass index (BMI), known to associate with T levels (28, 29), as a covariate in our GWAS. Including up to 127 covariates (based on (53)), e.g., assay center, dilution factors, blood draw time, and socioeconomic status indicators, or excluding related individuals from the analysis all showed negligible effects on the genetic findings we report here (Supplementary Figure 8).

Independent lead SNPs were selected for each chromosome by recursively taking the SNP with the lowest p-value (until none below the *p*-value threshold 5e-08 were left) from the GWAS summary statistics and removing all SNPs 500kb on each side of it from the next round. The chromosomal positions of these 1mb windows were stored, and overlapping windows were merged into the final list of loci. The SNP with the lowest p-value in each of these windows was selected as the lead SNP.

### Pathway, tissue enrichment and co-localisation analyses

Tissue and gene set enrichment analyses were carried out with SNP2GENE and GENE2FUNC implemented in FUMA using default settings (54). For testing in which tissues the genes residing in the GWAS loci were preferably expressed, we used the full distribution of SNP p-values and the GTEx v6 30 general tissue types. For pathway analysis, to assess whether the genes in the GWAS loci are overrepresented in pre-defined gene sets via hypergeometric tests, we selected manually curated KEGG-pathways (55). For co-localisation analyses to assess whether the genetic loci showed evidence for shared genetic effects between males and females, and to estimate the maximum posterior probability (MAP) for the loci being shared, we used gwas-pw (56).

### Replication in the Young Finns Cohort and calculation of PGS

The Cardiovascular Risk in Young Finns Study (YFS) is a longitudinal follow-up of 3,596 subjects at baseline. The baseline survey was conducted in 1980 and subsequent follow-ups involving the whole sample were held in 1983, 1986, 2001, 2007, 2011, and 2017. For testosterone and SHBG, we used data on 2001 follow-up (Subjects aged 24-39 yrs). A venous blood sample was drawn from antecubital vein after 12-hour overnight fast. Serum was aliquoted and stored in −70 Celsius degrees until analysis. In males, total testosterone quantification was performed in 2009 with competitive radioimmunoassay (Spectria Testosterone kit, Orion Diagnostica, Espoo, Finland) and Bio-Rad Lyphocheck control serums 1, 2, and 3 were used in quality control. Before quantification, serum aliquots had been melted three times. Total testosterone was quantified first and aliquots were re-frozen before SHBG quantification. In females, total testosterone quantification was performed in 2011. SHBG quantification for males was done in 2009 and for females in 2011 with Spectria SHBG IRMA kit (Orion Diagnostica, Espoo, Finland). Free testosterone was estimated using Vermeulen’s formula. As albumin concentration was not available, we used fixed albumin concentration of 43 g/l.

In YFS, genotyping was performed at the Wellcome Trust Sanger Institute (UK) using customized Illumina Human Map 670k bead array. The custom content on 670k array replaced some poor performing probes on Human610 and added more CNV content. As quality control, we excluded individuals and probes with over 5% of missingness (--geno and --mind filters in Plink). Variants deviating from Hardy-Weinberg equilibrium (p < 1×10^-6) and minor allele frequency below 1% were excluded. Related samples were excluded (n=51) with pi-hat cut-off of 0.2. Total of 2,442 individuals and 546,674 variants passed the quality control measures. The mean call rate across all included markers after the quality control was 0.9984. Next, imputation was performed using population-specific Sequencing Initiative Suomi (SISu) as reference panel. We examined the association of the UK Biobank GWAS lead SNPs with the corresponding T trait in YFS. If the annotated lead SNP was not available in YFS, we used LDstore2 (v2.0b) (57) to calculate LD in a 100kb window around the lead SNP in the UK Biobank imputed data and selected the closest SNP with R2>0.8 with the lead SNP as a proxy.

To construct PGS we applied the LDpred (28) method to the sex-specific GWAS results from the UK Biobank for total T, SHBG, FAI and free T using 1000 Genomes Europeans as LD reference and the default LD radius to account for LD. We then used the weights from the LDpred infinitesimal model to construct genome-wide PGSs for each individual in the YFS with Plink 2.0 (11 Feb 2018). Only variants imputed with high confidence (imputation INFO ≥ 0.8) were included in PGS calculation. Variants in chromosomes 1-22 and chrX were included. In males, allele dosage of 2 was used for X-chromosomal haploid variants. To evaluate the PGS prediction accuracy in the YFS, we calculated the R2 for each trait using using linear regression with z-score normalized PGS as predictor, age and 10 PCs as covariates and z-score normalized T trait as outcome.

### Estimation of heritability and calculation of the genetic correlations by LDSC

SNP-based heritability for the studied T traits and genetic correlations between these and with 44 additional phenotypes were estimated using linkage disequilibrium score regression (LDSC)(35). The summary statistics for the 44 traits were downloaded directly from the source repositories and analysed locally, for the original sources please see references in Supplementary Table 25. For the genetic correlation analyses, pre-computed LD Scores from 1000 Genomes Europeans excluding the HLA region were used. For 23 traits we performed the analyses using sex-specific GWAS results and compared these to data from sex-combined GWAS (Supplementary Table 27). Generally, genetic correlation results using either sex-specific or sex-combined GWAS data were highly similar.

### Disease associations in FinnGen

To assess if the PGS for studied traits associate with disease risk we utilized the FinnGen study (data freeze 5), consisting of 217,464 (94,478 males, 122,986 females) (29). FinnGen is comprised of Finnish prospective epidemiological and disease-based cohorts and voluntary biobank samples collected by hospital biobanks. The genotypes have been linked to national hospital discharge (available from 1968), death (1969–), cancer (1953–) and medication reimbursement (1964–) registries as well as the registry on medication purchases (1995-). The samples were genotyped with Illumina and Affymetrix arrays (Illumina Inc., San Diego, and Thermo Fisher Scientific, Santa Clara, CA, USA). The genotypes have been imputed with using the SISu v3 population-specific reference panel developed from high-quality data for 3,775 high-coverage (25-30x) whole-genome sequencing in Finns. The detailed genotype imputation workflow can be found at https://dx.doi.org/10.17504/protocols.io.xbgfijw. The dataset uses genome build 38 (hg38).

For PGS analyses, we used same variant weights (LDpred infinitesimal model) as for YFS, and calculated genome-wide PGSs for each individual with PLINK2 (v2.00a2.3LM). Variants in chromosomes 1-22 and chromosome X (imputed with high confidence, imputation INFO ≥0.7) were included (total number of variants ranging from 6,535,263 for female total T to 6,536,405 for female SHBG) and we used genotype dosages to incorporate imputation uncertainty. In males, allele dosage of 2 was used for X-chromosomal haploid variants. We studied the PGS associations to 36 disease endpoints with potential links to androgens, representing six loosely defined disease categories. For details of the studied phenotypes see Supplementary Table 21, www.finngen.fi and risteys.finngen.fi. Cox proportional hazards models were used for estimating hazard ratios (HRs) and 95% CIs, with age as the time scale and 10 first principal components of ancestry and genotyping batch as covariates. The proportionality assumption for Cox models was assessed with Schoenfeld residuals and log-log plots. For the cross-sex analyses, we took the sex-specific PGSs, and checked whether these would associate with the studied endpoints in the other sex, using the z-test to compare equality between the original and cross-sex associations. We additionally performed SHBG-adjusted PGS associations to all endpoints to control for potential confounding of SHBG to total and free T.

Patients and control participants in FinnGen provided informed consent for biobank research, based on the Finnish Biobank Act. Alternatively, older research cohorts, collected prior the start of FinnGen (in August 2017), were collected based on study-specific consents and later transferred to the Finnish biobanks after approval by Valvira, the National Supervisory Authority for Welfare and Health. Recruitment protocols followed the biobank protocols approved by Valvira. The Coordinating Ethics Committee of the Hospital District of Helsinki and Uusimaa (HUS) approved the FinnGen study protocol Nr HUS/990/2017.

The FinnGen study is approved by Finnish Institute for Health and Welfare (THL), approval number THL/2031/6.02.00/2017, amendments THL/1101/5.05.00/2017, THL/341/6.02.00/2018, THL/2222/6.02.00/2018, THL/283/6.02.00/2019, THL/1721/5.05.00/2019, Digital and population data service agency VRK43431/2017-3, VRK/6909/2018-3, VRK/4415/2019-3 the Social Insurance Institution (KELA) KELA 58/522/2017, KELA 131/522/2018, KELA 70/522/2019, KELA 98/522/2019, and Statistics Finland TK-53-1041-17. The Biobank Access Decisions for FinnGen samples and data utilized in FinnGen Data Freeze 6 include: THL Biobank BB2017_55, BB2017_111, BB2018_19, BB_2018_34, BB_2018_67, BB2018_71, BB2019_7, BB2019_8, BB2019_26, Finnish Red Cross Blood Service Biobank 7.12.2017, Helsinki Biobank HUS/359/2017, Auria Biobank AB17-5154, Biobank Borealis of Northern Finland_2017_1013, Biobank of Eastern Finland 1186/2018, Finnish Clinical Biobank Tampere MH0004, Central Finland Biobank 1-2017, and Terveystalo Biobank STB 2018001.

### Causality analyses

MR analyses treat genetic variants as instrumental variables and their reliability depends on two key assumptions: 1) alleles are randomly assigned, and 2) that alleles that influence exposure do not influence the outcome via any other means. The first assumption is controlled by using BOLT-LMM as our model in the primary GWAS analysis, but the second is harder to control when using a large number of SNPs as instrumental variables. In line with the observed wide-spread genetic pleiotropy affecting most complex traits, we noted that the GWAS loci contained many genes associated with pleiotropic effects on human phenotypes (for example, *LIN28B (58), GCKR (56)* and *TYK2(59)*). Therefore, given the vast polygenicity of the studied T traits, we chose latent causal variable (LCV) (31) and MR-Egger (32) as our primary MR methods, designed to take into account pleiotropy-induced confounding when assessing causal relationships. LCV has been proposed to provide more unbiased causality estimates than conventional MR approaches, whereas MR Egger should provide accurate causality estimates under the InSIDE assumption (the genetic variants have pleiotropic effects that are independent in magnitude and are thus not mediated by a single confounder exposure), besides its recommended use as a sensitivity analysis for conventional MR (31, 32). For comparison we also ran conventional MR analyses (Inverse-Variance Weighed (IVW)), that remains more sensitive for confounding by genetic correlation and pleiotropy (31). To extend the basic MR Egger analysis and to tease out the potential effects of SHBG on causality estimates of total and free T, we used multivariable MR Egger (33). LCV reports genetic causality proportion (GCP) as an estimate of causality, under a model where genetic correlation between two traits is mediated by a latent variable having a causal effect on each trait. GCP=1 means trait 1 is fully correlated with the latent variable, and hence fully causal to trait 2. A high GCP value and a statistically significant effect support partial genetic causality between the traits, and suggest that interventions targeting trait 1 are likely to affect trait 2. The *p*-value obtained in the analysis refers to the null hypothesis that the GCP=0. A highly significant *p*-value does not require a high GCP. Positive GCP value indicates causality of trait1 to trait2, whereas a negative value indicates support for causality of trait 2 to trait 1. LCV also estimates genetic correlation between the traits. To estimate the how much T could explain sex differences in hemoglobin we calculated ((FT_m-FT_f/SD_FT_m)*β*SD_H_m)/((H_m-H_f)/SD_H_m), where FT = mean free Testosterone, m=males, f=females, SD = standard deviation, β=MR estimate, H=Hemoglobin in UK Biobank based on (60). We applied the LCV, MR-Egger, multivariate MR-Egger and IWV models locally using R 4.0.2. The MR analyses were run using TwoSampleMR (v0.5.2) (61) and MendelianRandomization (v0.5.0) R packages (62). For the traits from public GWAS included in genetic correlation analyses, in 16 out 44 instances we could perform two sample MR (phenotype data not based on UK Biobank samples, Supplementary Table 24). FinnGen represents an independent research cohort from the UK Biobank and thus all FinnGen causality analyses were two-sample MR analyses.

## Data Availability

Data from FinnGen and the UK Biobank are available for researches by application. For YFS see https://youngfinnsstudy.utu.fi/index.html. Summary stats from publicly available GWAS can be downloaded from the source repositories for each study.

https://www.finngen.fi/en/access_results

https://www.ukbiobank.ac.uk/

## Acknowledgements / Funding statement

The research has been conducted using the UK Biobank Resource under application number 22627. The FinnGen project is funded by two grants from Business Finland (HUS 4685/31/2016 and UH 4386/31/2016) and eleven industry partners (AbbVie Inc, AstraZeneca UK Ltd, Biogen MA Inc, Celgene Corporation, Celgene International II Sarl, Genentech Inc, Merck Sharp & Dohme Corp, Pfizer Inc., GlaxoSmithKline, Sanofi, Maze Therapeutics Inc., Janssen Biotech Inc). The funders had no role in study design, data collection and analysis, decision to publish or preparation of the manuscript. The following biobanks are acknowledged for collecting the FinnGen project samples: Auria Biobank (https://www.auria.fi/biopankki), THL Biobank (https://thl.fi/fi/web/thl-biopankki), Helsinki Biobank (https://www.terveyskyla.fi/helsinginbiopankki), Biobank Borealis of Northern Finland (https://www.oulu.fi/university/node/38474), Finnish Clinical Biobank Tampere (https://www.tays.fi/en-US/Research_and_development/Finnish_Clinical_Biobank_Tampere), Biobank of Eastern Finland (https://ita-suomenbiopankki.fi), Central Finland Biobank (https://www.ksshp.fi/fi-FI/Potilaalle/Biopankki), Finnish Red Cross Blood Service Biobank (https://www.veripalvelu.fi/verenluovutus/biopankkitoiminta) and Terveystalo Biobank (https://www.terveystalo.com/fi/Yritystietoa/Terveystalo-Biopankki/Biopankki/). All Finnish Biobanks are members of BBMRI.fi infrastructure (www.bbmri.fi). The Young Finns Study has been financially supported by the Academy of Finland: grants 322098 (T.L), 286284, 134309 (Eye), 126925, 121584, 124282, 129378 (Salve), 117787 (Gendi), and 41071 (Skidi); the Social Insurance Institution of Finland; Competitive State Research Financing of the Expert Responsibility area of Kuopio, Tampere and Turku University Hospitals (grant X51001); Juho Vainio Foundation; Paavo Nurmi Foundation; Finnish Foundation for Cardiovascular Research; Finnish Cultural Foundation; The Sigrid Juselius Foundation; Tampere Tuberculosis Foundation; Emil Aaltonen Foundation; Yrjo Jahnsson Foundation; Signe and Ane Gyllenberg Foundation; Diabetes Research Foundation of Finnish Diabetes Association; This project has received funding from the European Union’s Horizon 2020 research and innovation programme under grant agreements No 848146 for To Aition and grant agreement 755320 for TAXINOMISIS; European Research Council (grant 742927 for MULTIEPIGEN project); Tampere University Hospital Supporting Foundation and Finnish Society of Clinical Chemistry. We greatly thank all UK Biobank, the Young Finns study, and FinnGen participants, as well as the principal investigators, laboratory personnel and data management teams behind these efforts. This work was supported by the Academy of Finland (https://www.aka.fi/en/) grants 331671 to N.M., 312072 to Tm.T., 288509 and 319181 to M.P., and 315589 and 320129 to T.T., and Academy of Finland Center of Excellence in Complex Disease Genetics grants 312075 to M.D., 312062 and 336820 to S.R. and 312076 to M.P.), by the Finnish Foundation for Cardiovascular Research (https://www.sydantutkimussaatio.fi/en) (S.R.), the Sigrid Juselius Foundation (https://sigridjuselius.fi/en/) (S.R., M.P., T.T.) and University of Helsinki (https://www.helsinki.fi/en) HiLIFE Fellow and Grand Challenge grants (S.R. and M.P.), and three-year research project grant (T.T.).

