## Supplementary Figures and Legends for "Genetic analyses on the health impacts of testosterone highlight effects on female-specific diseases and sex differences"

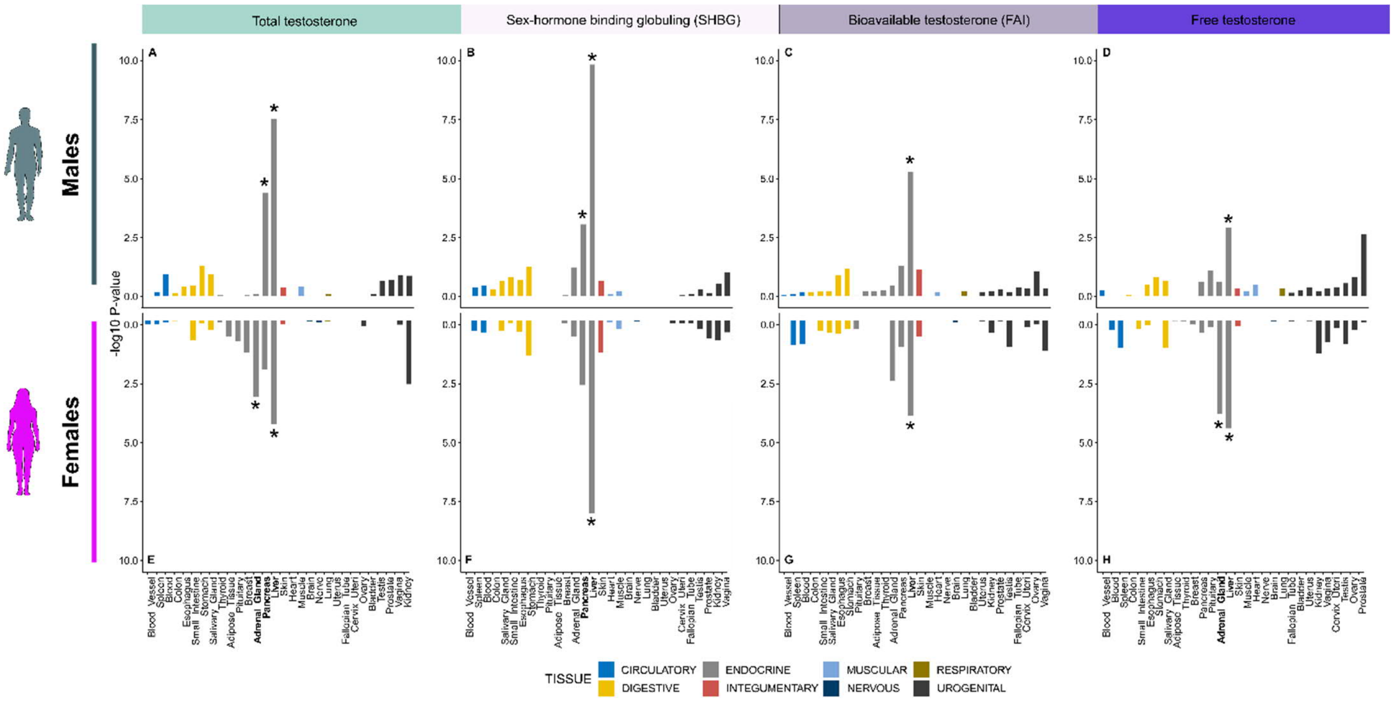


Supplementary Figure 1. Results from the tissue enrichment analyses from MAGMA implemented in FUMA. Panels A-D show tissue enrichment in males, and E-H in females. The GTEx tissues show in figure have been group based on tissue types. For all the studied testosterone traits, the genes residing in the associated loci based on the UK Biobank GWAS are preferably expressed in the liver. We detected no evidence for the GWAS loci being enriched for genes expressed in the testis or the ovaries, potentially pointing to T metabolism rather than T production having larger effects on adult serum T levels. However, consistent with the notion that the adrenal gland is a major site for T production in females, many genes in the loci associated with total and free T in females appeared preferably expressed in this tissue. Likely underscoring the close relationship between metabolism and T levels, pancreas also shows up as a tissue enriched for the GWAS signals for total T and SHBG in males.

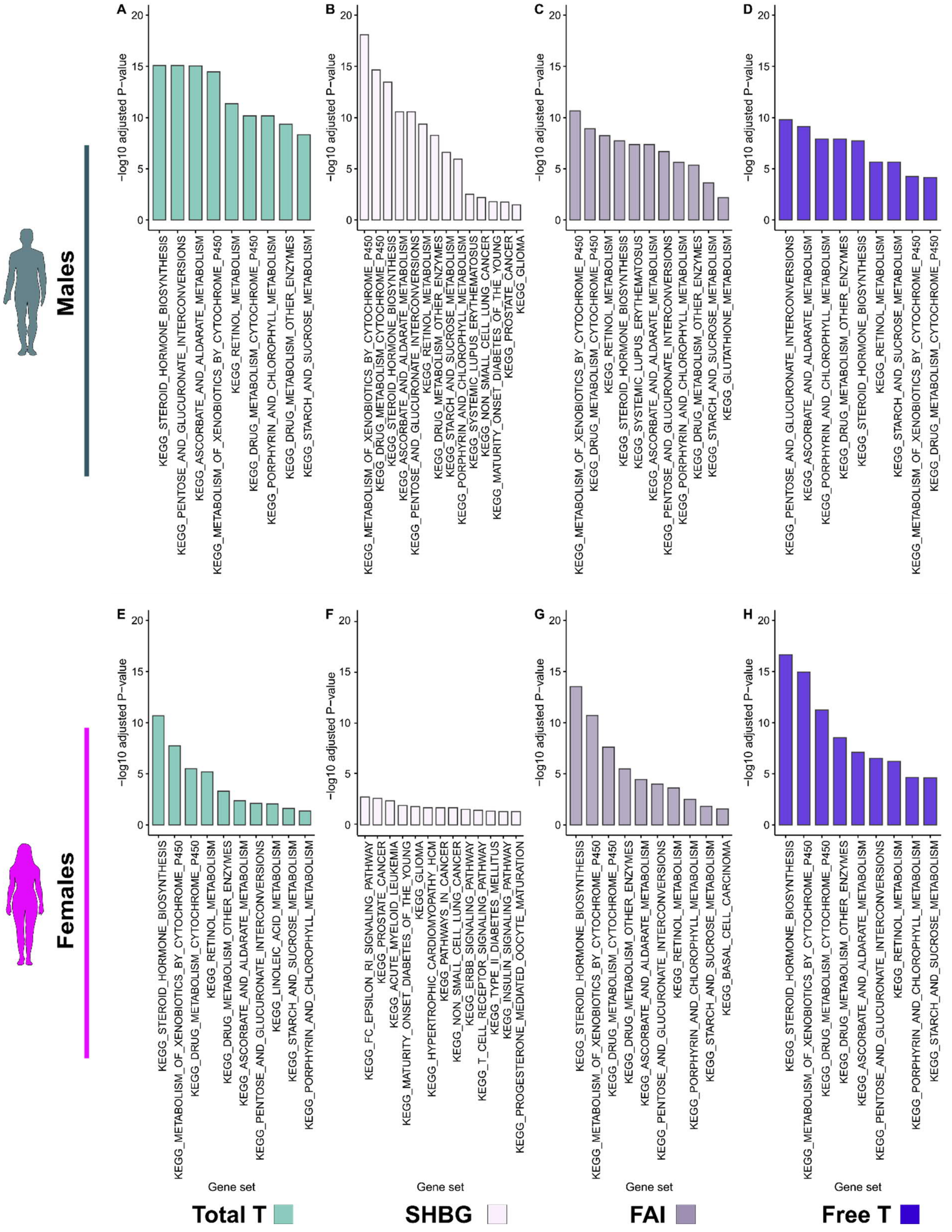


Supplementary Figure 2. Visualisation of the results from KEGG pathway analyses from FUMA. The figure illustrates Kyoto Encyclopedia of Genes and Genomes (KEGG) pathways showing enrichment for the genes residing in the GWAS loci. The GWAS loci were enriched for genes that affect steroid hormone biosynthesis, metabolism and excretion, with the exception of SHBG in females. A) Total T males; B) SHBG males; C) FAI males; D) Free T males; E) Total T females; F)

SHBG females, G) FAI females; H) Free T, females. T=Testosterone, SHBG = Sex-hormone binding globulin, FAI = Free androgen index, Free T = calculated free testosterone = Free T. Results shown for all KEGG pathways with *p* for enrichment < 0.05 after adjusting for multiple testing in FUMA.

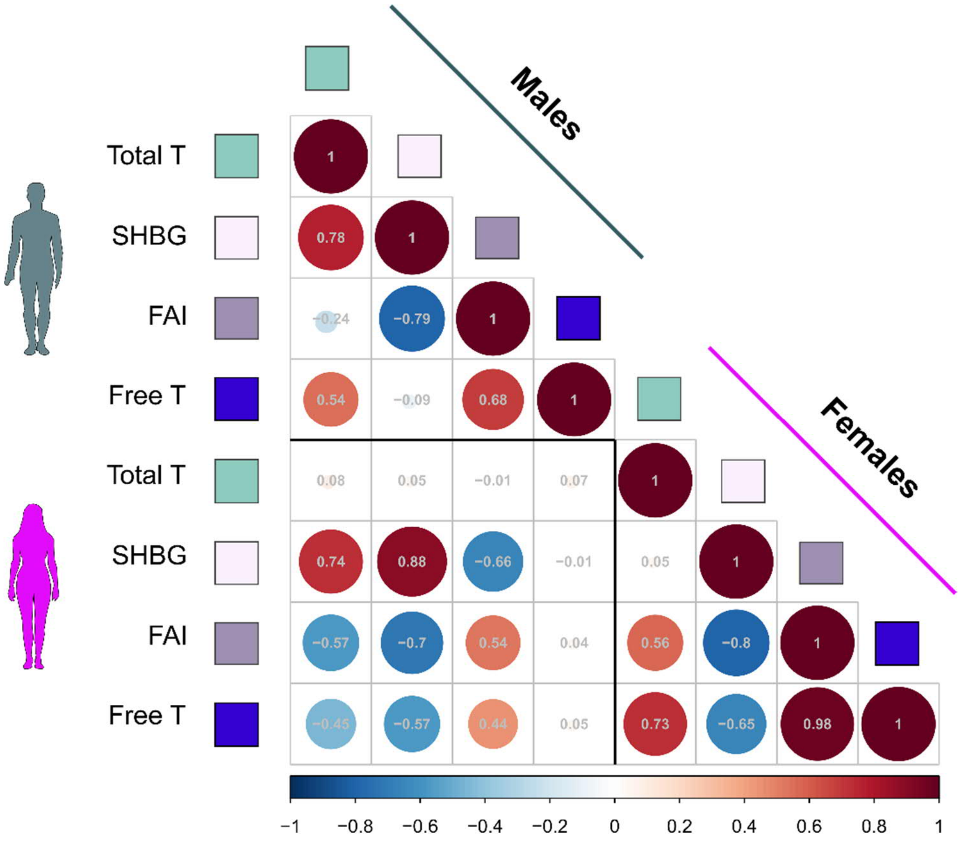


Supplementary Figure 3. Genetic correlation results between the studied testosterone traits. Correlations are shown for males, females and between the sexes, based on LDSC analyses. Generally, the genetic factors affecting total T in females and free T in males appeared unconnected to any other studied traits in the opposite sex. In line with recent findings, we observed a weak genetic correlation for total T levels between males and females ($r_g$=0.08, *p*=0.063). Similarly, free T showed a low genetic correlation estimate between the sexes ($r_g$=0.05, *p*=0.102), whereas the genetic correlation for SHBG was high ($r_g$=0.88, *p*=9.7e-197), and for FAI intermediate ($r_g$=0.57, *p*=5.8e-26). Moreover, consistent with the epidemiological observations that total T and SHBG levels are correlated in males, these traits showed a positive genetic correlation ($r_g$=0.78, *p*=1.0e-127). Yet, for females such connection did not exist ($r_g$=0.05, *p*=0.207).

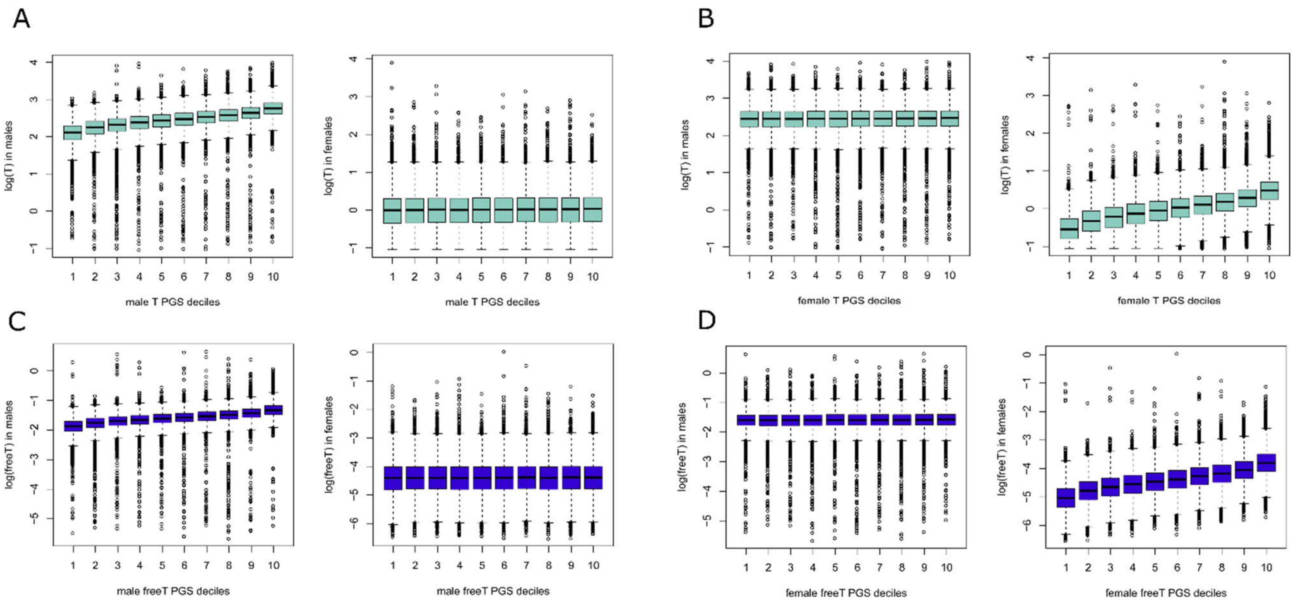


Supplementary Figure 4. Predictive ability of total T and free T PGSs for T levels in the UK Biobank. The figure shows how the sex-specific PGS correlates with T levels only within the sex it was constructed, and has no predictive ability in the opposite sex. Although the cross-sex comparisons in UK Biobank are performed using independent datasets (males vs. females), note the training and the test datasets for the sex-specific PGSs include the same individuals, leading to inflated results. A) Male Total T PGS, B) Female Total T PGS, C) Male Free T PGS, D) Female Free T PGS. The box plots show median (black line), lower and upper quartiles (colored area of the box) for log T and free T in PGS deciles, and the error bars indicate 5% and 95% quantiles.

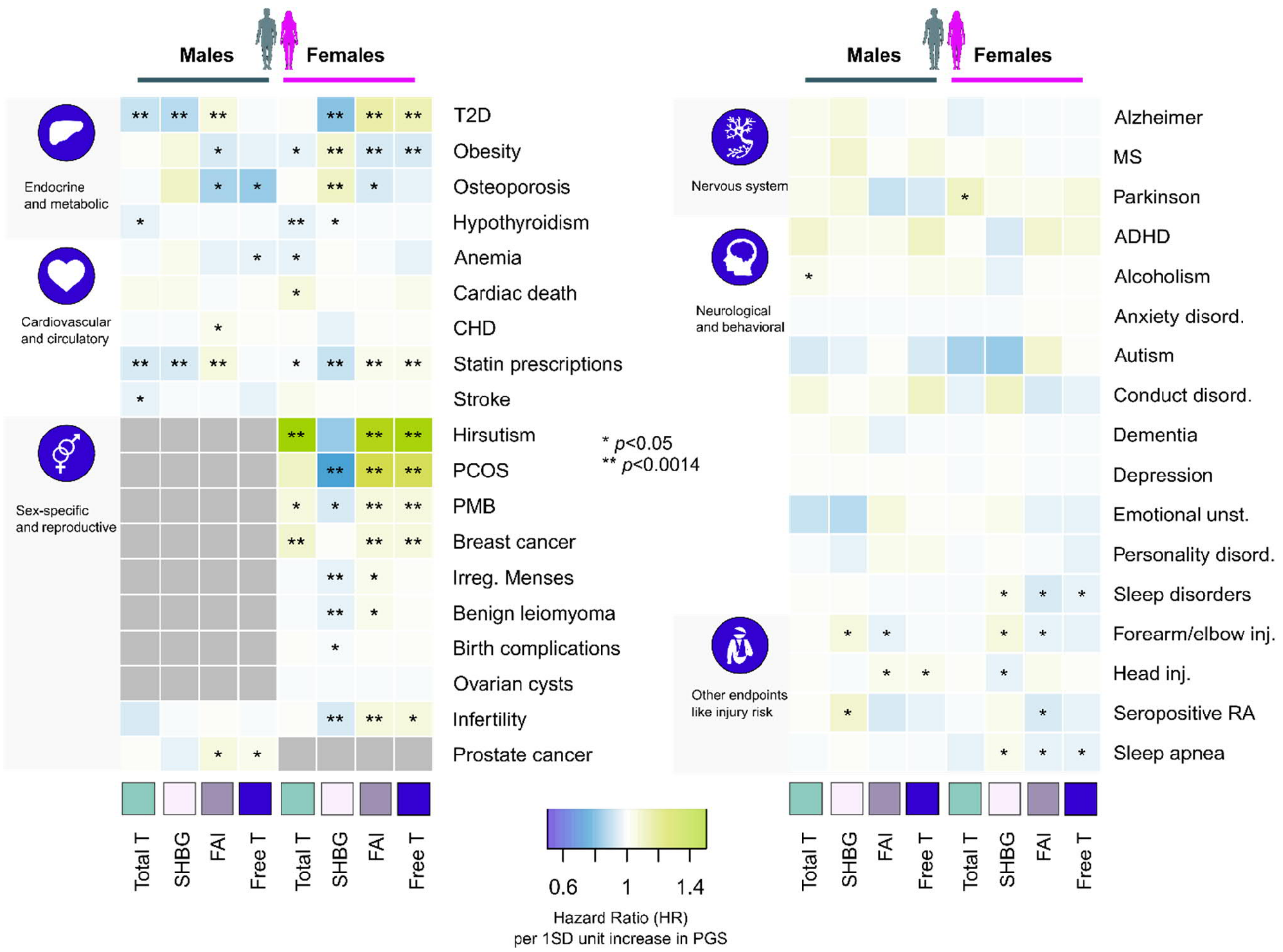


Supplementary Figure 5. PGS associations in the FinnGen. The heatmap illustrates hazard ratios (HR) per 1SD increase in PGS for all studied traits in the sex-specific analyses. *p*<0.0014 corresponds to Bonferroni correction for 36 traits.

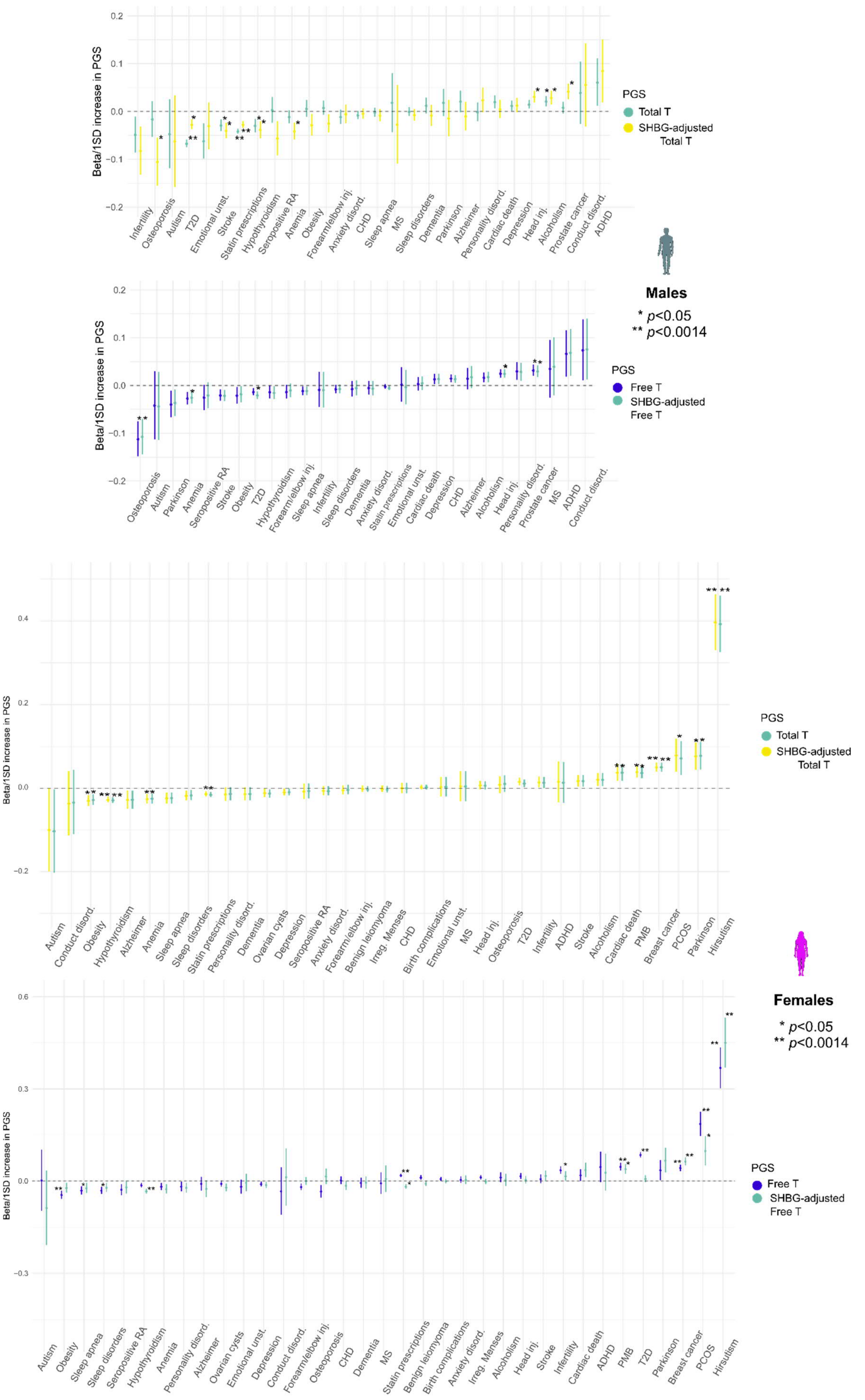
Beta/1SD increase in PGS
PGS
Total T
SHBG-adjusted Total T
Free T
SHBG-adjusted Free T
Males
* p<0.05
** p<0.0014
Females
* p<0.05
** p<0.0014
Infertility
Osteoporosis
Autism
T2D
Emotional unst.
Stroke
Statin prescriptions
Hypothyroidism
Seropositive RA
Anemia
Obesity
Forearm/elbow inj.
Anxiety disord.
CHD
Sleep apnea
MS
Sleep disorders
Dementia
Parkinson
Alzheimer
Personality disord.
Cardiac death
Depression
Head inj.
Alcoholism
Prostate cancer
Conduct disord.
ADHD
Conduct disord.
Ovarian cysts
Benign leiomyoma
Irreg. Menses
Birth complications
PMB
Breast cancer
PCOS
Hirsutism

Supplementary Figure 6. Comparison of unadjusted and SHBG-adjusted T PGS associations in FinnGen. The figure shows disease risk in standardised units per 1SD increase in PGS. For some traits with known T contribution the effects between the original unadjusted and the adjusted analysis appear different (e.g. SHBG-adjusted total T but not unadjusted total T showing nominally significant association to osteoporosis in males, but surprisingly also PCOS signals being lost after SHBG adjustment). For most other traits, the effects estimates remain highly similar between the unadjusted and adjusted analyses. Yet notably the total T PGS association to T2D vanishes in both sexes when taking into account the effect of SHBG in the adjusted analysis.

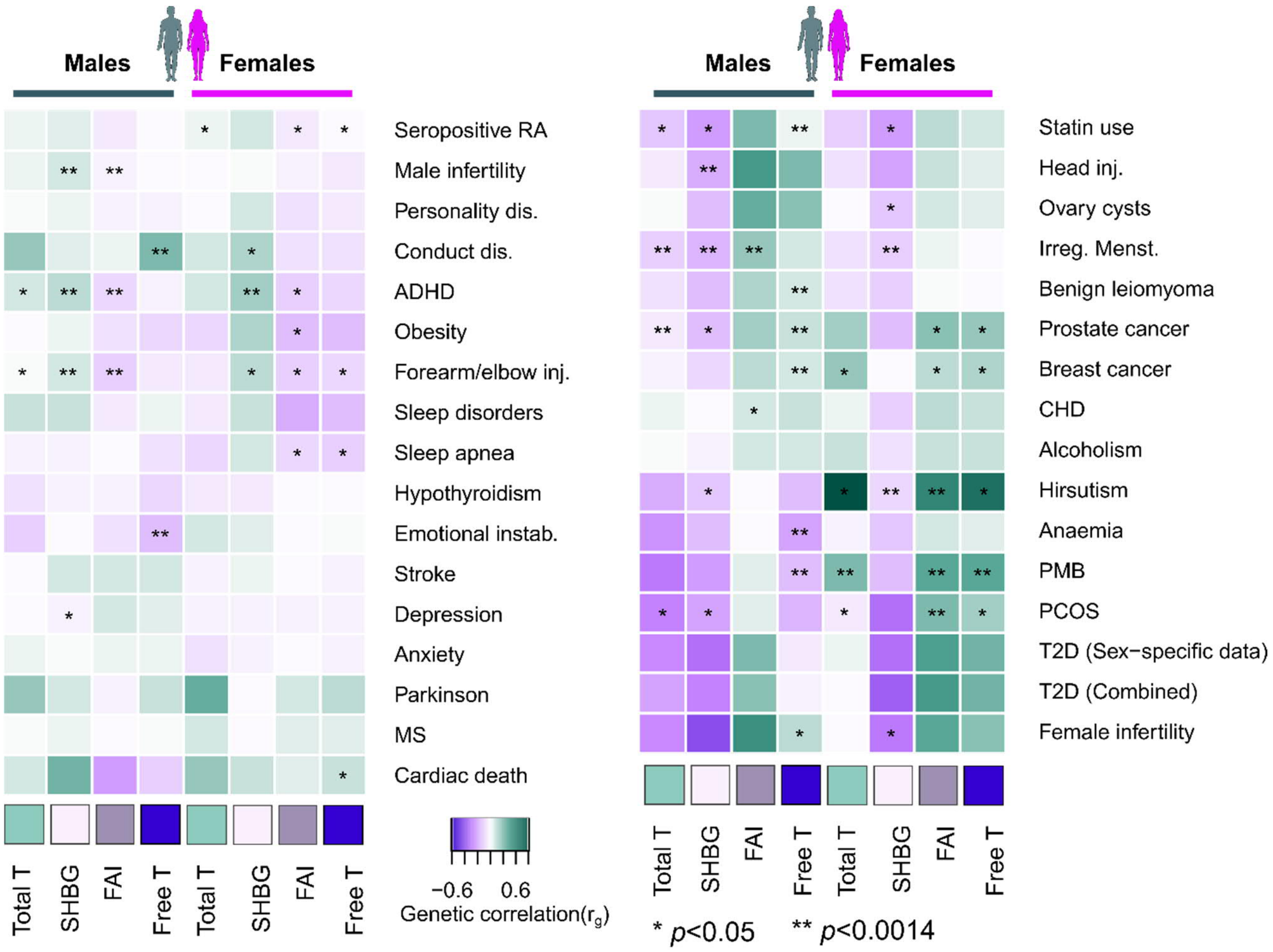


Supplementary Figure 7. Visualisation of genetic correlations and causality analyses between T, SHBG, FAI and free T and the studied disease endpoints in FinnGen. Heatmap colors indicate the direction and the strength of the genetic correlation (purple, negative correlation, green, positive correlation) based on LCV. Asterisks indicate statistical support (either $p$<0.05 or $p$<0.0014) for causality between traits based on either LCV or MR Egger analyses (Supplementary Table 22).

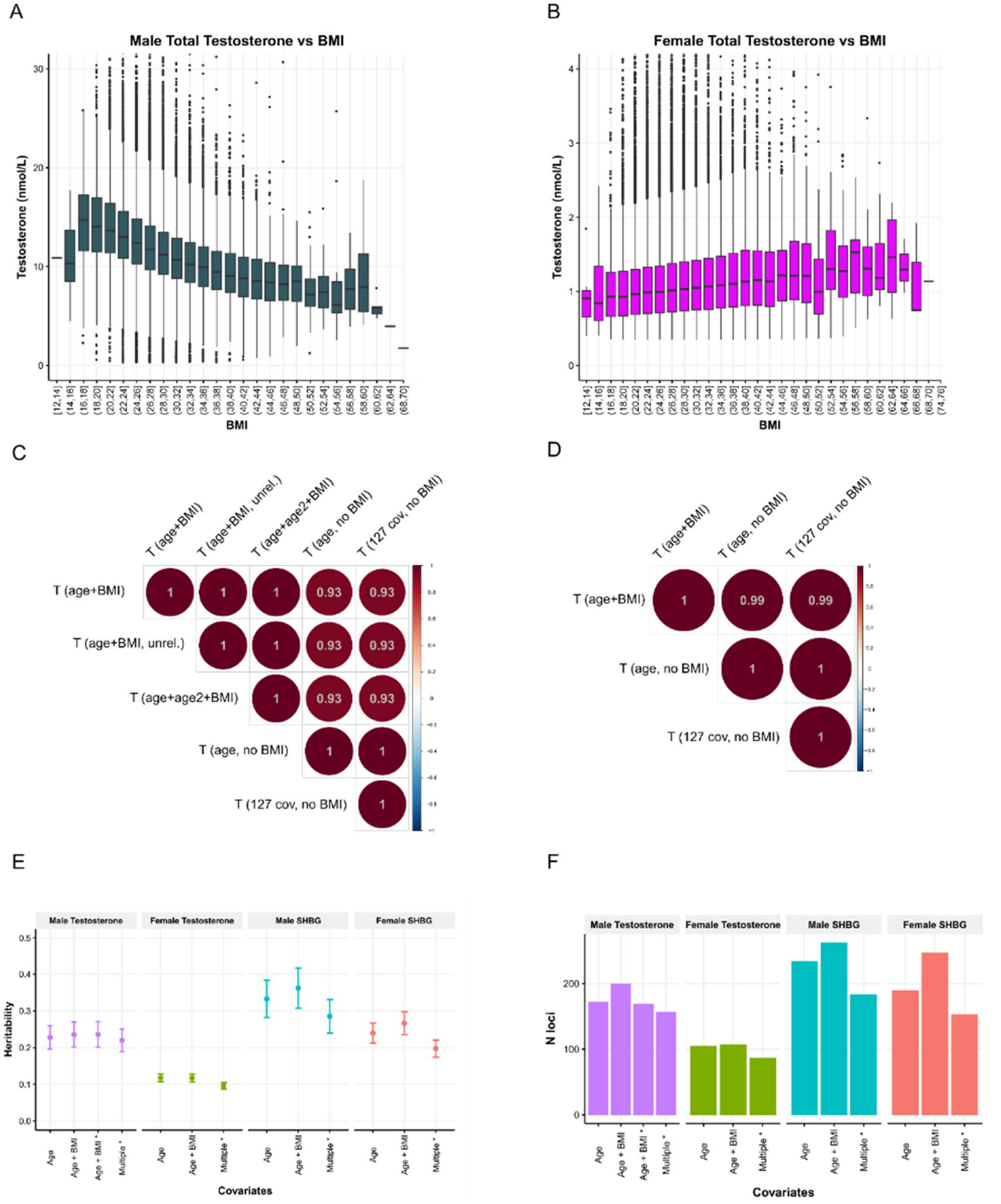


Supplementary Figure 8. The effects of body mass index (BMI) on testosterone (T) levels and GWAS results in the UK Biobank. Panels A and B show the relationship between raw total T levels and BMI (binned in two unit intervals) in the UK Biobank for males (A) and females (B). In males, a significant negative correlation exists between BMI and T levels (r=-0.30, *p*=0), whereas this relationship is reversed in females (r=0.08, *p*=2.9e-230). Panels C and D show genetic correlations from LDSC between total T GWAS runs under varying covariate combinations in males (C) and females (D), implying BMI has the largest effects on results. Adding BMI as a covariate increased both the estimated heritability (LDSC) and the number of loci found for both total T and SHBG (E and F). The other 127 tested covariates based on Sinnott-Armstrong et al. (2019) with sample dilution factors included, show negligible effects on the results in genetic correlation analyses.
